# Universal HIV screening: perspectives from Eastern Romania

**DOI:** 10.64898/2026.08.19.26360868

**Authors:** Cabiria M Barbosu, Carmen Manciuc, Timothy Dye

## Abstract

HIV/AIDS remains a major global public health challenge, and disparities in HIV testing persist. In 2024, an estimated 87% of people living with HIV were aware of their status, with lower testing coverage among children aged 0–14 years (63%), and among men (84%) compared with women (92%). Romania initiated its national AIDS program in 1995 and quickly progressed in addressing the epidemic; however, HIV testing remains largely concentrated in specialized services, with late diagnosis, missed testing opportunities, and stigma continuing to limit timely identification and linkage to care. This study aimed to understand better HIV testing/screening practices among clinicians in eastern Romania and to identify gaps that could be addressed through medical education. We conducted an analytical cross-sectional study among healthcare providers in the eastern region of Romania to assess whether HIV testing is routinely offered to patients, explore gaps in clinical judgment and perceived responsibility, and identify factors that facilitate HIV testing. A 17-question anonymous survey was distributed via WhatsApp to clinician groups between August 1 and September 30, 2023. Respondents included physicians (71.9%), nurses (28.1%), and other healthcare professionals, working in infectious diseases (36.0%), internal medicine (22.3%), primary care (13.7%), and other specialties, such as obstetrics-gynecology and pediatrics (18.0%). Only 38.1% of respondents reported routinely screening all patients aged 18 years and older for HIV, while 61.9% did not offer regular HIV testing. The most cited reasons for not screening were the perception that HIV testing was not their responsibility and that their department did not require testing (18.1% each). Clinicians working in settings with established policies on HIV confidentiality, non-discrimination, testing, and post-exposure prophylaxis were more likely to offer routine testing. Universal HIV screening remains uncommon among clinicians in eastern Romania. Supportive institutional policies appear to facilitate routine testing and may reduce missed opportunities for early diagnosis. Normalizing HIV testing as part of routine clinical care, in line with the Romanian National Health Strategy 2022–2030, is crucial for enhancing early detection and strengthening prevention efforts through coordinated action among clinicians, public institutions, and civil society.

## Introduction

Globally, HIV/AIDS remains a significant public health issue, with 40.8 million people living with HIV in 2024, even though new HIV infections have been reduced by 61% since the peak in 1996. [1] The 2025 UNAIDS goal was to reach 95-95-95 targets for HIV testing, treatment, and viral suppression. However, disparities in HIV testing remain. In 2024, only 87% of people living with HIV knew their status, and children aged 0-14 years are less likely to be tested (63%), compared with men (84%) and women (92%). [2] As of July 2025, in Eastern Europe and Central Asia, about 2.1 million people are living with HIV, with 130,000 new infections in adults over 15 years, and 1,200 new infections in children aged 0-14 years. [2]

Romania, located at the crossroads of Central, Eastern, and Southeastern Europe, has a unique HIV infection history defined by a large pediatric epidemic during 1980-1990, with the leading cause of infection being the reuse of needles and syringes in hospitals and orphanages, combined with flawed medical practice, and the refusal of the government in power to acknowledge that HIV infection was present in the country. [3, 4] Another notable characteristic was the high prevalence of subtype F-1 strains causing infections, which differ from those typically found in Central/Eastern Europe. [5]

Romania launched its AIDS program in 1995, moving quickly to address the epidemic. [4] Romania’s HIV testing strategy follows the same public health approach recommended by the European Center for Disease Prevention and Control (ECDC): expanding access to testing, including community models, integrating testing into antenatal and clinical services, ensuring linkage to care, and monitoring late diagnoses. However, testing remains concentrated in specialized service settings, and late diagnosis and stigma persist. [6, 7]

HIV infection in Romania spiked since the beginning of the SARS-CoV-2 pandemic, with people newly diagnosed with HIV characterized by opportunistic infection and late diagnosis; in fact, approximately one-third of people newly diagnosed with HIV already have advanced disease. [8] As such, our study aimed to deepen the understanding of HIV testing and screening practices among clinicians in eastern Romania.

## Methods

We conducted an analytical cross-sectional study among healthcare providers in the eastern region of Romania (Moldova) to assess whether clinicians regularly offer HIV testing, identify gaps in clinical judgment, and determine areas where targeted medical education is needed. The survey consisted of 17 questions and was distributed between August 1 and September 30, 2023, with advertisements appearing three times.

The recruitment announcement, which included a link to an anonymous Google Docs survey and a corresponding QR Code, was distributed to a convenience sample of healthcare providers recruited via WhatsApp groups. Eligible participants included physicians, dentists, nurses, social workers, and pharmacists. Our prior research has shown that social media, particularly Facebook and WhatsApp, is an effective modality for reaching providers, with 2/3 or more of Romanian clinical providers reporting daily or weekly use of these platforms. [9]

### Outcome

This self-reported routine clinical practice analysis’s primary outcome is whether the clinicians offer HIV testing to all patients over the age of 18. We asked:

#### Do you offer HIV testing to all your patients over the age of 18? (Yes/No)

Healthcare practitioners who answered ’No’ were asked follow-up questions about the reasons for not testing and suggestions on what would facilitate their offering tests.

### Statistical analysis

Bivariate statistical analyses were conducted to establish associations between variables and the outcome measure of interest, using the Pearson chi-square test. Statistical significance was defined as the asymptotic significance (two-sided) of the chi-square value, set at p < 0.05. For multivariate modeling of the outcome of interest, we included all variables that were at least marginally associated with the outcome at the bivariate level. We then used the backward elimination procedure to remove variables from the multivariate model. [10] We used 0.25 as the cutoff p-value for entry into the model and 0.10 as the threshold for remaining in the model. [11] We assessed the overall significance of the resulting multivariate model at p < 0.05 and evaluated the model fit using the Pearson chi-square goodness-of-fit test and p-value. An appropriate fit model was defined as one with a p-value greater than 0.05. For bivariate analyses, we used logistic regression-derived Odds Ratios (OR) and their corresponding 95% confidence intervals to denote the magnitude of association. We used adjusted Odds Ratios (aOR) to indicate the same from multivariate analyses. SPSS 29.0.2.0 (IBM Corporation, Armock, NY, USA) was used for analyses.

The project was approved by the Research Subjects Review Board at the University of Rochester and the Ethics Committee of the “Sf Paraschiva” Clinical Hospital for Infectious Diseases in Iasi, Romania (decision no. 6/06.26.23). The University of Rochester’s Research Subjects Review Board office determined that this project met federal and university criteria for exemption (STUDY00008788). The project was conducted in accordance with the ethical standards outlined in the 1964 Declaration of Helsinki and its subsequent amendments.

## Results

Shown in Table 1, only 38.1% of participants reported screening all patients aged 18 and older for HIV. The most common reasons provided for not screening included “*It is not my responsibility*” and “*The department I work in does not require testing*” (18.1% in each category). 12.2% of clinicians did not consider their patients at risk, 10.8% indicated they were uncomfortable offering HIV testing unless there was a good reason to do so, and 9.4% expressed a need for additional HIV testing training. When asked what would help them recommend testing to all eligible patients, 36.0% wanted more justification for the importance of testing, 17.3% wanted training on discussing screening with patients, and 14.4% wanted more training on testing.

**Table 1.**
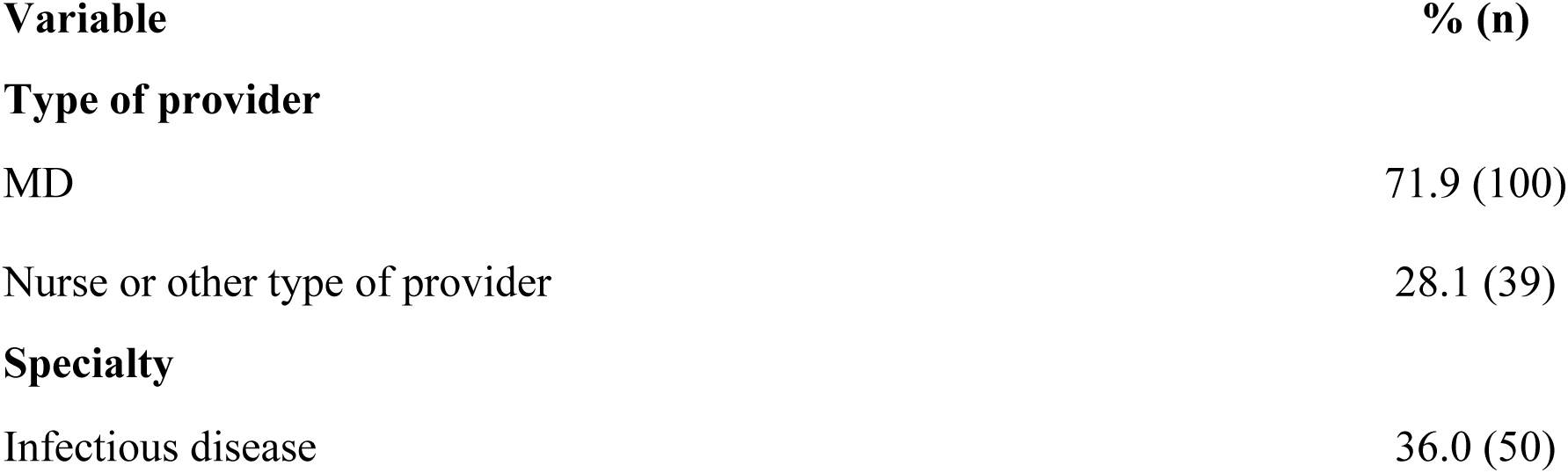
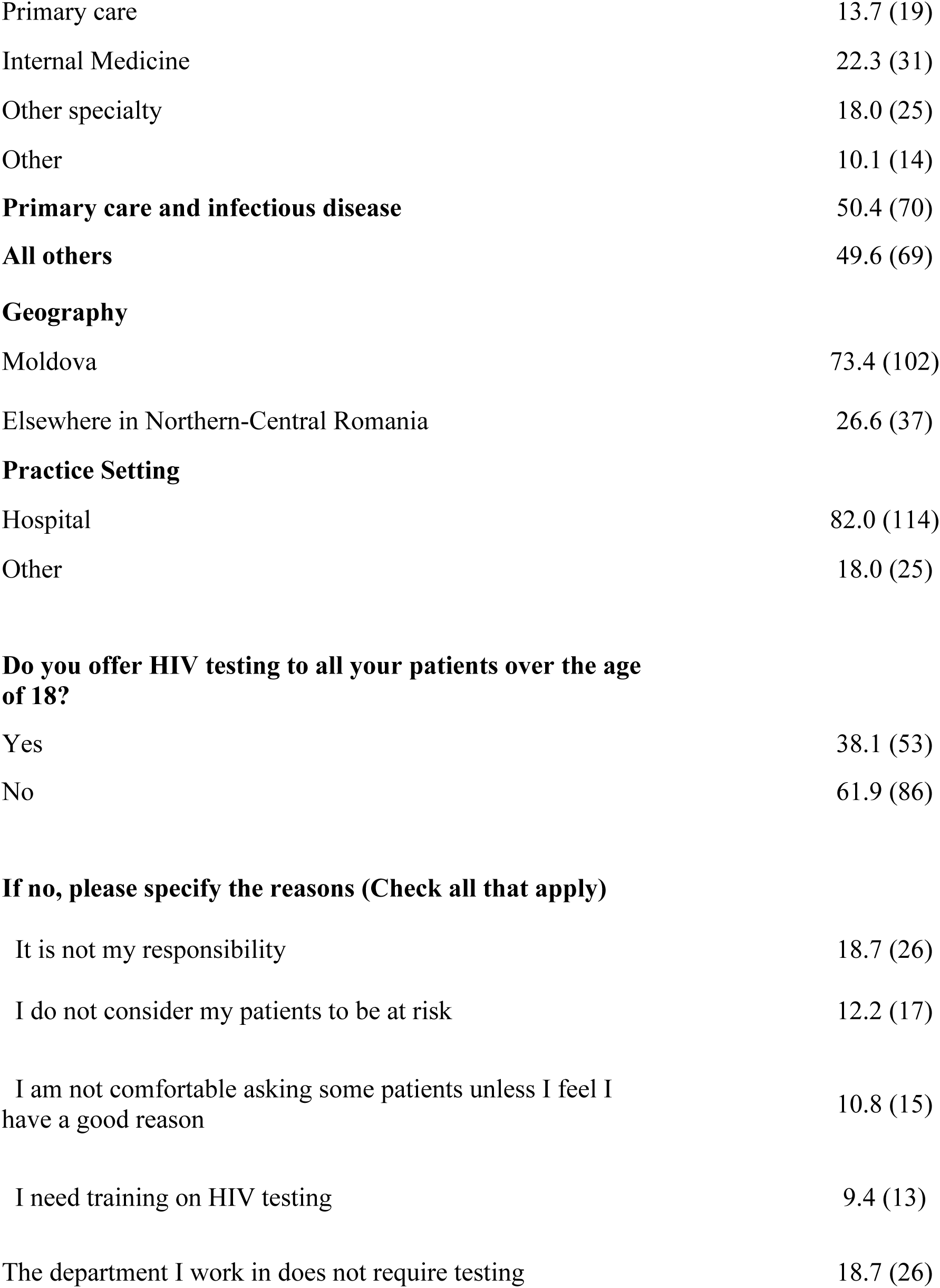
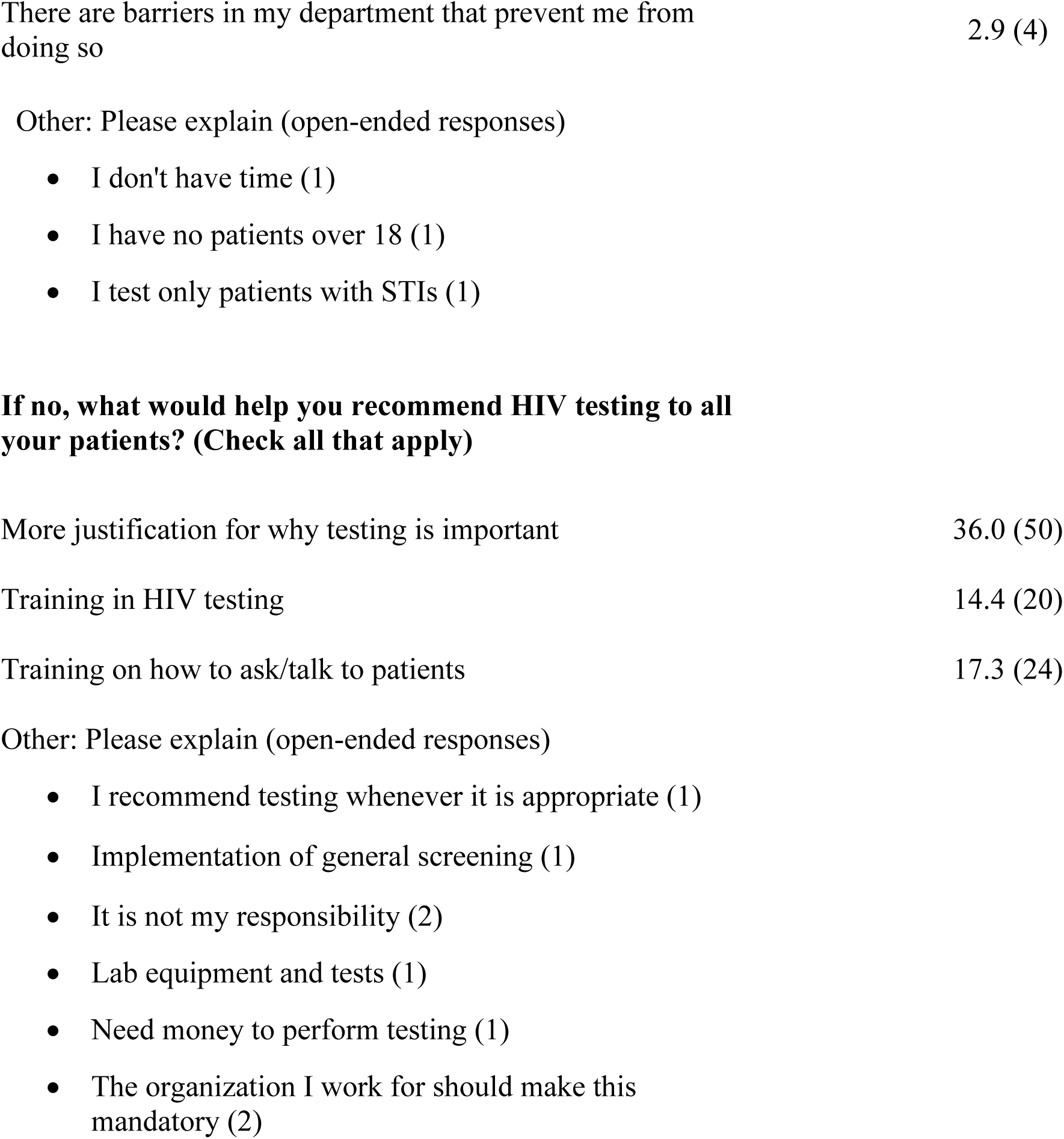
Participant, practice characteristics, and HIV universal screening.

As seen in Table 2, physicians were more likely (77.4%) to offer an HIV test. By specialty, infectious disease specialists offered the most HIV testing (54.9%), followed by internal medicine specialists (17%), and only 5.7% of primary care physicians offered it.

**Table 2.**
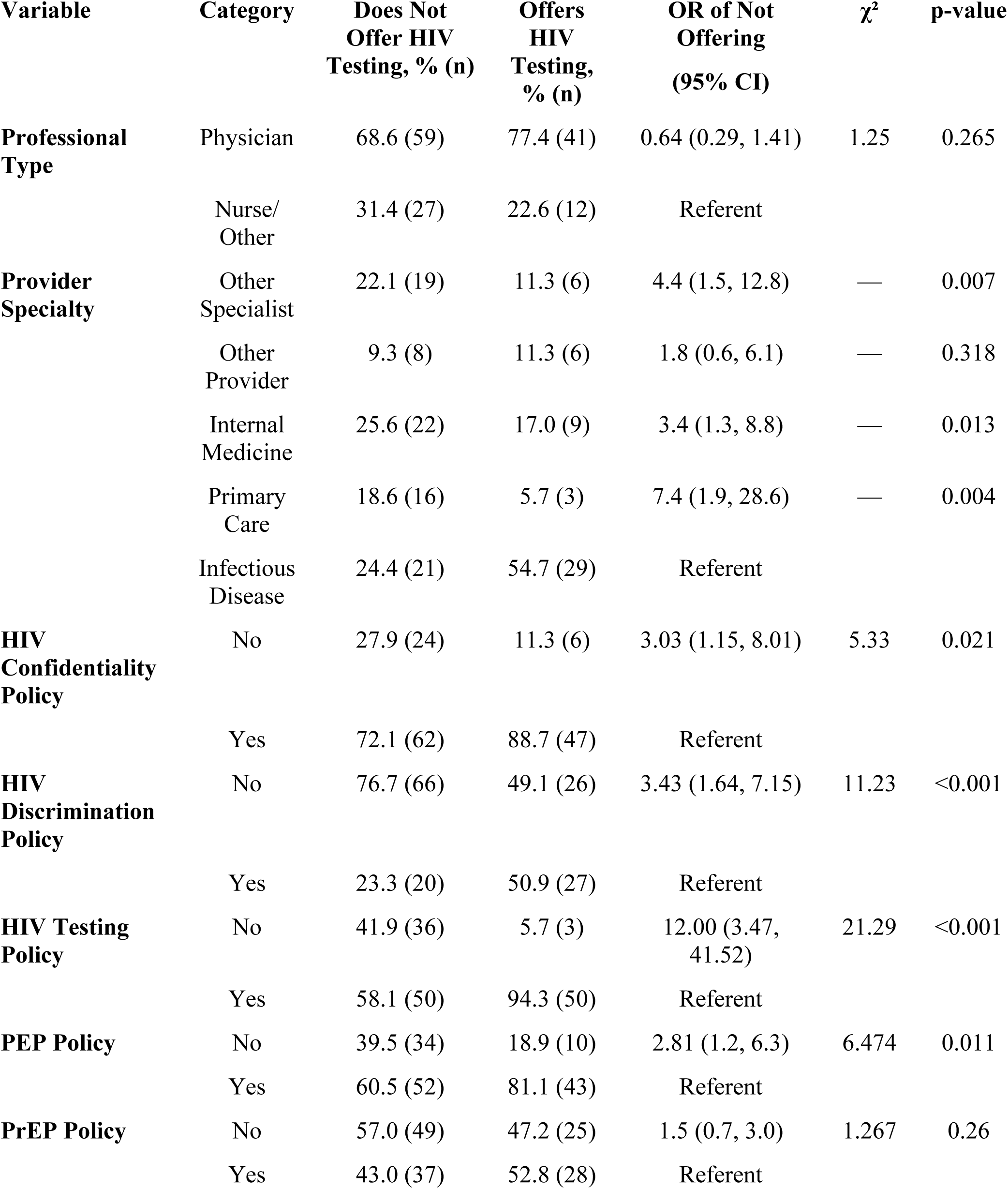
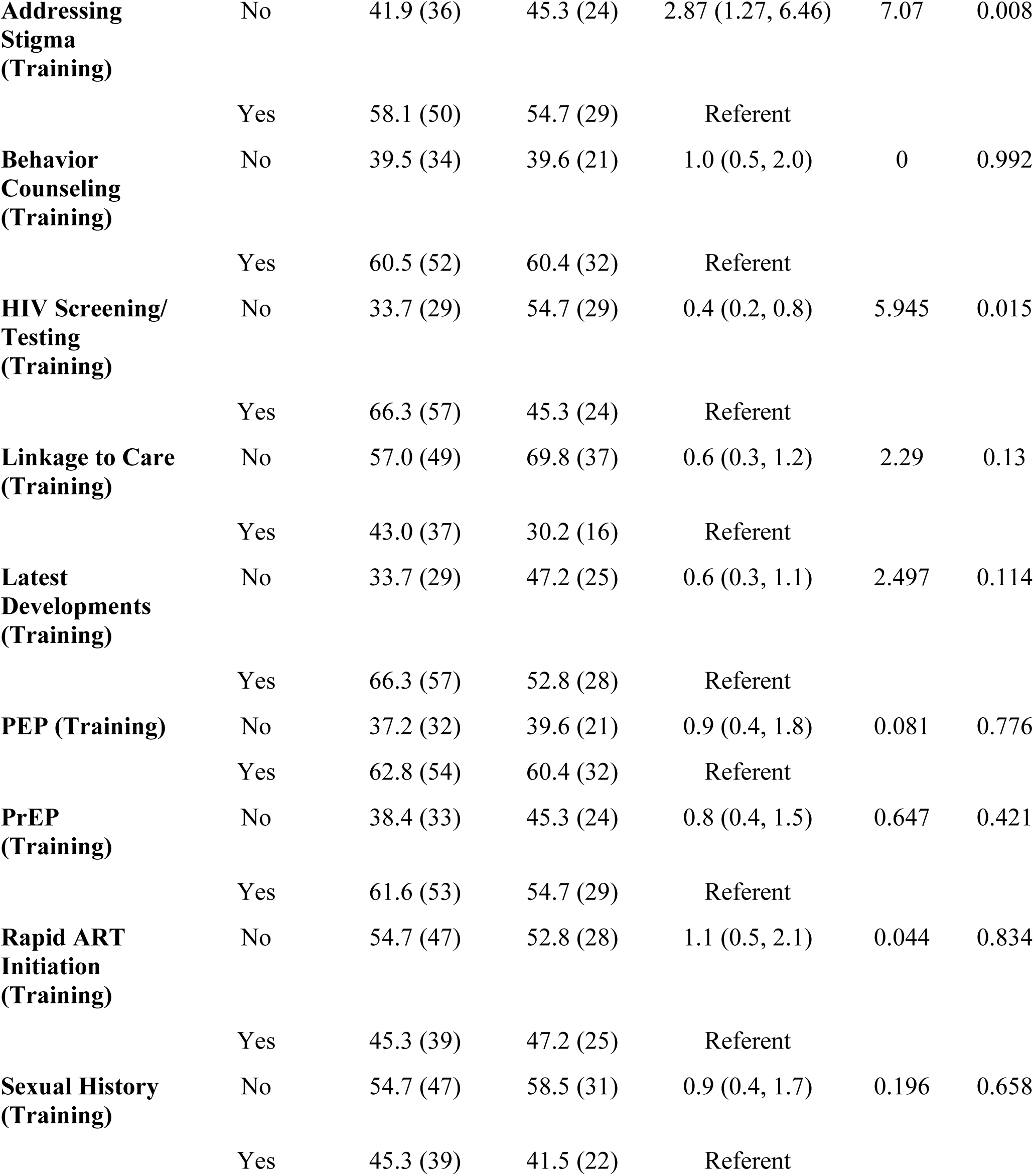
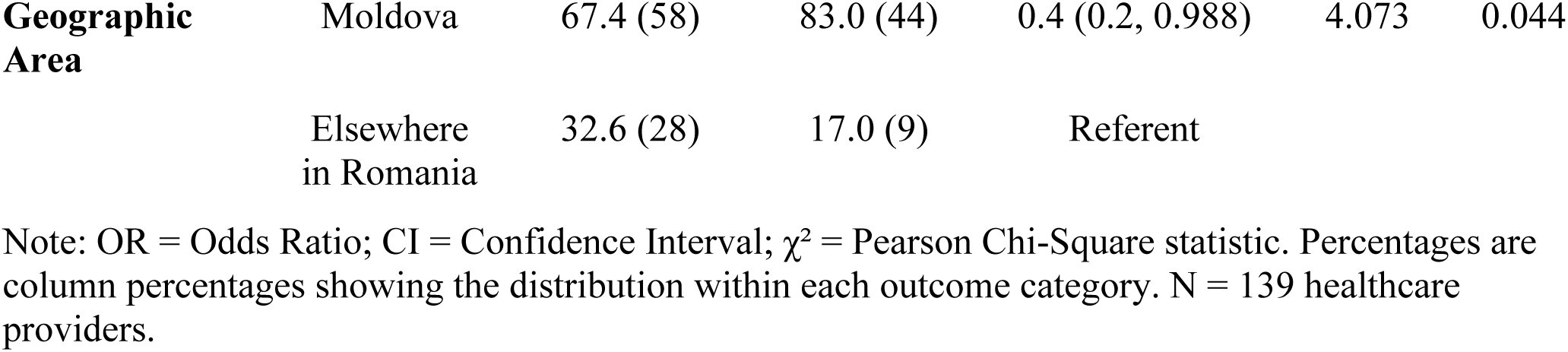
Bivariate Analysis of Factors Associated with Offering HIV Testing to Patients Over 18 Years Old.

As detailed in Table 3, after controlling for confounding effects, having no written HIV testing policy in their workplace was significantly associated with not offering screening to all patients over 18 years old, and not having an HIV discrimination policy was marginally associated. Healthcare practitioners not offering universal screening were significantly more likely to request training in screening/testing and in the latest HIV/AIDS developments.

**Table 3.** Multivariate Logistic Regression Model of Factors Associated with Not Offering HIV Testing to Patients Over 18 Years Old.

| Variable | B | SE | Wald | p-value | aOR | 95% CI for aOR |
| --- | --- | --- | --- | --- | --- | --- |
| **HIV Discrimination Policy** |  |  |  |  |  |  |
| No | 0.856 | 0.449 | 3.632 | 0.057 | 2.4 | 1.0–5.7 |
| Yes (ref) | — | — | — | — | — | — |
| **HIV Testing Policy** |  |  |  |  |  |  |
| No | 2.898 | 0.694 | 17.438 | <0.001 | 18.1 | 4.7–70.7 |
| Yes (ref) | — | — | — | — | — | — |
| **HIV Screening/Testing Training** |  |  |  |  |  |  |
| Not selected | −1.051 | 0.468 | 5.041 | 0.025 | 0.3 | 0.1–0.9 |
| **Selected (ref)** | — | — | — | — | — | — |
| **Latest Developments Training** |  |  |  |  |  |  |
| **Not selected** | −0.947 | 0.451 | 4.402 | 0.036 | 0.4 | 0.2–0.9 |
| **Selected (ref)** | — | — | — | — | — | — |
| **Geographic Area** |  |  |  |  |  |  |
| **Moldova** | −1.474 | 0.54 | 7.437 | 0.006 | 0.2 | 0.08–0.7 |
| **Elsewhere (ref)** | — | — | — | — | — | — |

Model Fit Statistics
| **Statistic** | **Value** |
| --- | --- |
| **−2 Log Likelihood**<br>**(Final Model)** | **119.138** |
| **Likelihood Ratio Test**<br>**$\chi^2$** | **49.351** |
| **Likelihood Ratio Test**<br>**df** | **5** |
| **Likelihood Ratio Test**<br>**p-value** | **<0.001** |
| **Pearson Goodness-of-Fit**<br>**$\chi^2$** | **95.725** |
| **Pearson Goodness-of-Fit**<br>**p-value** | **0.814** |
| **Nagelkerke pseudo-R-square** | **0.406** |

**Notes:** aOR: Adjusted Odds Ratio. 95% CI: 95% confidence interval. Reference category for outcome: Yes (offers HIV testing). The final model was derived using a backward elimination stepwise selection method. The following variables were removed during stepwise selection as non-significant: Addressing Stigma ( $p = 0.807$ ), HIV Confidentiality Policy ( $p = 0.678$ ), Linkage to Care Options ( $p = 0.563$ ), Provider Specialty ( $p = 0.432$ ), and PEP Policy ( $p = 0.233$ ). $N = 139$ healthcare providers. aOR values $>1$ indicate increased odds of not offering HIV testing; values $<1$ indicate decreased odds.

## Discussion

We found that universal testing for HIV among practitioners in Romania is inconsistent, and among some practitioners, uncommon. Since 2018, the European Centre for Disease Prevention and Control (ECDC) has provided a framework to develop, implement, monitor, and evaluate HIV testing, emphasizing the need to (i) reach populations at higher risk of infection, (ii) diversifying testing services beyond traditional clinical settings; (iii) integrating HIV testing with other services (e.g., Hepatitis B & C) to improve uptake. [12] While ECDC recommendations should be applied throughout the region, national-level implementation varies. In some countries, for instance, there was a mandatory “offer” of HIV testing to all pregnant women; in others, testing is offered as part of routine antenatal tests (opt-out). In many countries, the approach is “opt-in,” requiring specific informed consent. Moreover, pre- and post-test counseling, partner notification, ARV treatment of mother and child, and follow-ups vary across countries. [13]

A 2022 systematic guideline review shows that national medical guidelines lack adequate testing recommendations, with fewer than half of national clinical guidelines for HIV indicator conditions in 15 European countries recommending HIV testing despite evidence. [13]

The first step to diagnose an infection is to test. However, some national clinical guidelines do not include HIV testing recommendations even when the condition is an “HIV indicator condition.” This omission is a missed opportunity to initiate treatment and prevent the spread of infection. [13] Variations in countries’ policies, such as opt-in vs. opt-out, counseling level, consent requirements, stigma, or access to self-testing, are factors resulting in uneven HIV testing and care access across Europe.

Our survey indicates that universal HIV screening in Moldova, Romania is uncommon, with only 38.1% of healthcare practitioners offering a test. Although most practitioners do not offer an HIV test to all patients aged 18 and older, our data suggest that having supportive written policies in the workplace can promote universal screening. However, many healthcare facilities lack such policies. Healthcare practitioners who do not offer universal screening expressed interest in HIV training, particularly on the rationale and importance of testing, and on how to conduct testing-related conversations with patients.

The International Association of Providers of AIDS Care (IAPAC) and several HIV community partners in Europe have raised concerns about achieving the 2025 (95-95-95) targets, noting that, despite frameworks, many European countries are not on track to meet them. They emphasized the need for a renewed 2016 EU Action Plan on HIV, Viral Hepatitis, TB, and other STIs, and the development of more effective policies for highly impacted communities, to end HIV/AIDS as a public health threat by 2030. [14] ECDC/WHO sets standards and recommendations; however, it is each Member State’s responsibility to set specific policies for its health system.

Our 2017 prior work conducted in Romania showed that 40.8% of the clinical providers did not provide HIV screening and testing to at least one segment of the population, with the most common reason being that they didn’t feel that screening was their responsibility (33.3%) and required more justification for why testing is important (27.5%). [15] The results of our present study indicate that HIV screening patterns are similar to those observed in our previous study (2017). However, direct comparisons are limited by methodological differences, suggesting the need for system-level interventions to address training and practice needs and sustain routine screening. A substantial prevention obstacle is that HIV testing remains concentrated in the specialized testing centers, hospitals/infectious disease clinics. NGOs and community services offer HIV testing in larger cities, but the scale of this service is limited. [16] Testing is offered routinely as part of prenatal care, included in the differential diagnosis, or performed by request. However, it is not routinely performed in family doctors’ offices, as some providers do not offer it for various reasons. [16, 17]

Romania’s national HIV testing data show that young adults aged 20–49 years constitute most new HIV diagnoses, with a notable gap between the age distribution of new diagnoses and the age groups seeking HIV testing, with younger individuals underrepresented in testing services. [18] This shows the need for targeted HIV testing initiatives aimed at increasing awareness and testing uptake among younger populations.

In Europe, HIV testing policies vary by country, although the ECDC recommends routine testing for people aged 15 - 49 and those over 50, and that testing should be offered to those under 15 and over 65 who are at increased risk. [12] In Romania, there is no official age range for HIV testing, and the law requires parental consent for minors (under 18) to access HIV testing and receive information about their status. The updated Romanian National Strategy for 2022-2030 does not specify a specific HIV testing age, emphasizing broad access and efforts that focus on high-risk populations. [19]

## Conclusions

The 2022–2030 Romanian National Health Strategy aims to modernize and expand public health services, including HIV care, to ensure accessible, well-coordinated care across all levels. [20] Increasingly, Romania recognizes that effective HIV management involves not only viral suppression but also addressing stigma, social inclusion, mental health, and quality of life for people living with HIV. [21] Achieving these objectives requires tackling geographic and regional disparities, securing sustainable funding, integrating medical, psychosocial, and social support services, and implementing robust monitoring and evaluation systems. [22] The strategy clearly identifies PrEP as a priority intervention. Initially, the plan aimed to provide PrEP to approximately 150 individuals through the national ART program by the end of 2024. [19]

In parallel, non-governmental organizations started offering PrEP in select centers; for example, ARAS’s “PrEPpoint” initiative serves men who have sex with men (MSM) and transgender individuals and reportedly reached about 220 people by 2024. [23] Despite its inclusion in the national strategy, PrEP availability remains limited, and the current targets appear modest, indicating a need for expansion. Additionally, the new long-acting injectable regimen (Vocabria + Rekambys) has received regulatory authorization in the European Union and is expected to be adopted in Romania as well. [24]

Despite widespread international efforts toward HIV eradication, the reality shows that this is a challenging task. Clinicians should normalize HIV testing as part of routine care to avoid missed opportunities for early diagnosis and to reduce stigma.

Study limitation: Because this study relied on self-reports, convenience sampling, and social media recruitment in Eastern Romania, the findings may not generalize widely and may be subject to selection bias.

## Data Availability

The data is available from the corresponding author upon reasonable request.

## Acknowledgments

Not applicable

